# Symptomatology during seasonal coronavirus infections in children is associated with viral and bacterial co-detection

**DOI:** 10.1101/2020.03.24.20038828

**Authors:** Emma M. de Koff, Marlies A. Van Houten, Elisabeth A.M. Sanders, Debby Bogaert

## Abstract

Lower respiratory tract symptoms during seasonal coronavirus infections in children are associated with RSV co-detection and increased levels of Haemophilus and Fusobacterium species.

## To the Editor

In the current SARS-CoV-2 pandemic (COVID-19), children seem to be less severely affected than adults. Case series of Chinese children with COVID-19 report mostly mild symptoms of a respiratory tract infection (RTI) [1, 2], though severe cases were also reported [3]. Children may however shed large quantities of viral particles in respiratory secretions and faeces, even when they are without symptoms of infection, and may thus contribute substantially to human-to-human transmission [4–6]. Coronavirus (HCoV) detection in asymptomatic children is quite common for other serotypes (OC43, NL63, 229E and HKU1) [7]. In case of severe HCoV-associated disease, younger age and chronic illness are identified risk factors [7, 8]. Furthermore, in HCoV-infected children, other respiratory viruses are often co-detected, and these other viruses may contribute to symptomatic illness [7]. Until now, bacterial co-infections have not been investigated. Improved understanding of determinants of symptomatic HCoV-associated illness may also increase our understanding of symptomatology and the potential role of transmission of COVID-19 by the paediatric population in the current pandemic. Because of a lack of data regarding the potential role of viral and bacterial co-infection in symptomatology and severity of HCoV infections and COVID-19, we evaluated respiratory viruses and the nasopharyngeal bacterial microbiome in the context of seasonal HCoV infections in a recent case-control study of children admitted to the hospital with lower respiratory tract infection (LRTI) in The Netherlands.

Between 2013 and 2015, we enrolled 154 children admitted with LTRI and aged between 4 weeks and 5 years, and 307 age-, gender- and season-matched healthy controls. Methods were previously published in detail [9]. The study protocol (www.trialregister.nl, NTR5132) was approved by the Dutch National Ethics Committee. Written informed parental consent was obtained from all participants. In the present work, we investigated whether symptomatology of HCoV infection is associated with viral and/or bacterial co-infections.

In short, nasopharyngeal swabs from cases at hospital admission and from healthy community-dwelling controls were tested for respiratory viruses using qualitative multiplex Real Time-PCR (RespiFinder® SMARTfast 22; Pathofinder, Maastricht, Netherlands) [10]. In addition, bacterial DNA was isolated and sequenced using 16S rRNA gene sequencing. Sequencing data is publicly available from the Sequence Read Archive database (BioProject ID PRJNA428382). Statistical analysis was performed in R (version 3.6.1). A p-value <0.05 or a Benjamini-Hochberg adjusted p-value <0.10 was considered statistically significant. Cases and controls were stratified according to HCoV detection (OC43, HKU1, NL63 or 229E). Differences in age were assessed with t-tests and differences in gender with chi-square tests. To test differences in viral (co-)detection and total number of viruses between groups, logistic and linear regression were used, respectively, adjusting for age. Differential abundances of the 50 most highly abundant operational taxonomic units (OTUs) were tested using *metagenomeSeq*, adjusted for age [11].

Viral data was available from 150 cases and 303 controls. HCoV was detected at a similar rate in 24 (16.0%) cases and in 58 (19.1%) controls (p=0.49). In all HCoV-positive children, both cases and controls, HCoV-OC43 was most commonly detected (62.5% cases, 51.7% controls; p=0.52), followed by HCoV-NL63 (16.7% cases, 27.6% controls; p=0.44), HCoV-229E (16.7% cases, 10.3% controls; p=0.67) and HCoV-HKU1 (12.5% cases, 12.1% controls; p=1.00). There was one case with co-detection of HCoV-OC43 and HCoV-229E, another case with both HCoV-OC43 and HCoV-HKU1, and one control with co-detection of HCoV-NL63 and HCoV-HKU1. HCoV was most frequently detected in the winter months, with 95.8% of HCoV-positive cases and 84.6% of HCoV-positive controls detected between December and March. Age was comparable between HCoV-positive cases (mean 17.0 months (SD 15.9)) and HCoV-positive controls (mean 16.4 months (SD 14.9), p=0.88), and between HCoV-positive and HCoV-negative cases (mean 17.5 months (SD 15.0), p=0.87). Gender was also comparable between HCoV-positive cases (50.0%) and HCoV-positive controls (41.4%; p=0.64) as well as HCoV-negative cases (36,6%; p=0.32).

Co-detection of other respiratory viruses with HCoV was more frequent in cases (96%) than in controls (69.0%; p=0.019) (Figure 1A). The total number of viruses detected in HCoV-positive cases amounted to maximum four (mean 2.5 (SD 0.9)), and was significantly higher than in HCoV-negative cases (mean 1.4 (SD 0.7); p<0.001) as well as in HCoV-positive controls (mean 2.1 (SD 0.9); p=0.043). Co-infection mostly concerned respiratory syncytial virus (RSV), which was even significantly more often detected in HCoV-positive cases (70.8%), than in HCoV-negative cases (43.9%; p=0.014; Figure 1A-B). By contrast, detection of rhinovirus was lower in HCoV-positive cases (37.5%) than in HCoV-positive controls (62.1%; p=0.042; Figure 1A). Co-detection of other viruses in HCoV-positive cases concerned adenovirus (16.7%), bocavirus (16.7%), human metapneumovirus (8.3%) and influenza virus (4.2%), though not different from HCoV-positive controls and HCoV-negative cases (Figure 1A-B).

**Figure 1.**
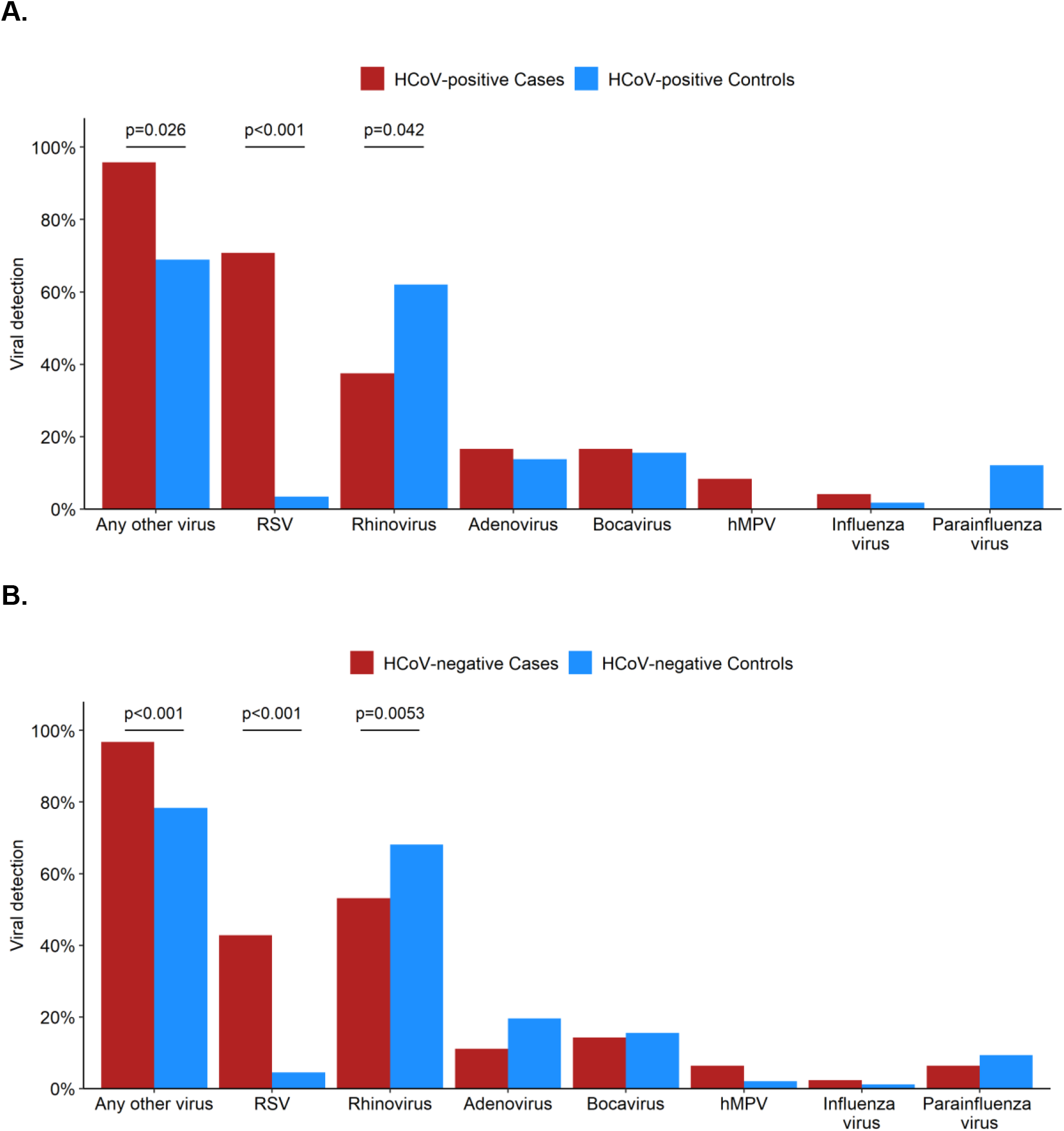
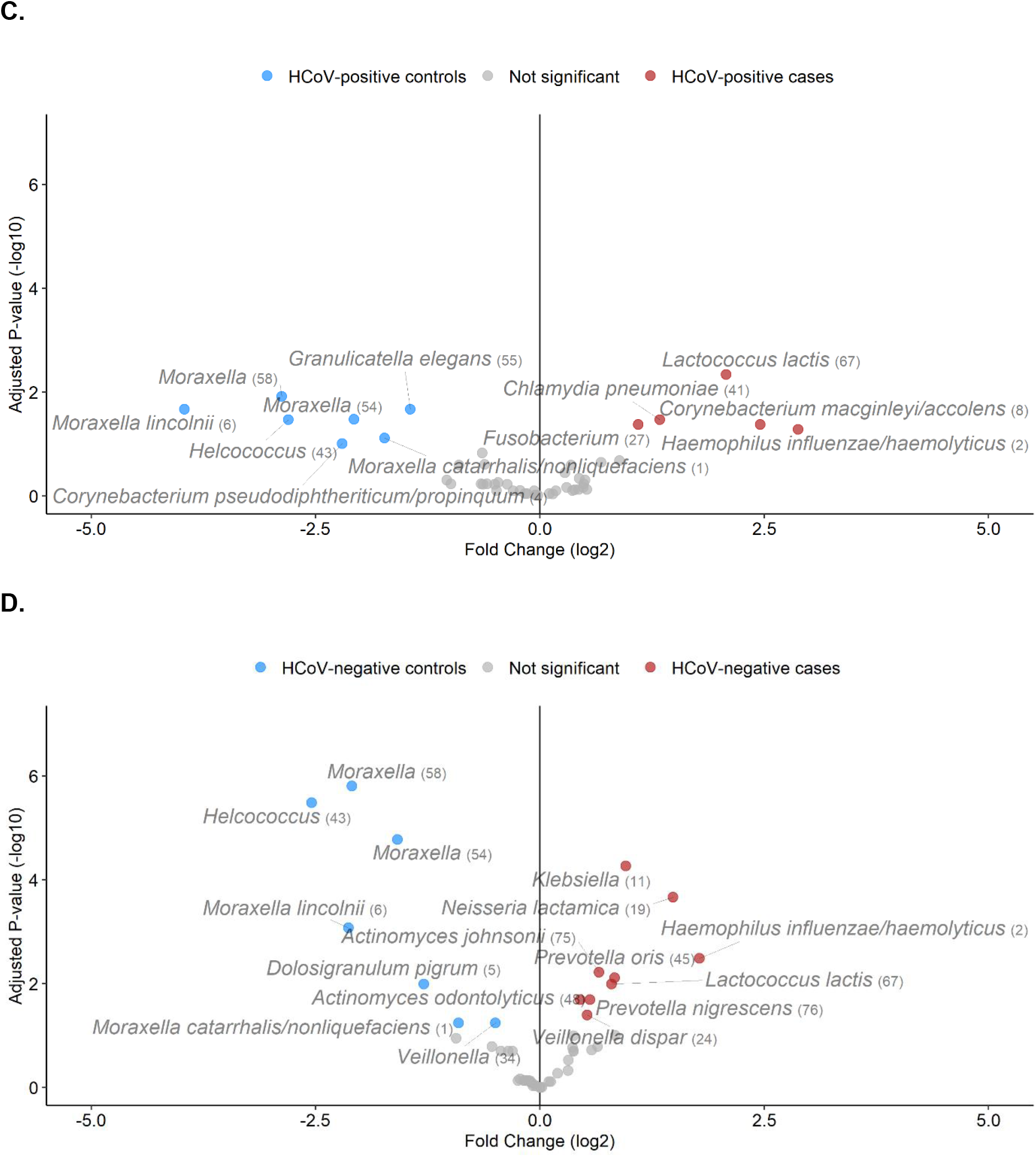
Viral and bacterial co-detection in cases and controls. Bar graphs (**A-B**) show viruses detected with qualitative PCR in **A**. coronavirus (HCoV-)positive cases and HCoV-positive controls and **B**. HCoV-negative cases and HCoV-negative controls. Vulcano plots (**C-D**) show log2 fold changes and -log10-transformed Benjamini-Hocberg adjusted p-values for differentially abundant bacterial taxa between **C**. HCoV-positive cases and HCoV-positive controls and **D**. HCoV-negative cases and HCoV-negative controls. Labeled dots represent OTUs that were significantly increased (*metagenomeSeq*, corrected for age, adjusted p-value <0.1) in either cases (red) or controls (blue). Any other virus = any respiratory virus that is not HCoV; RSV = respiratory syncytial virus; hMPV = human metapneumovirus.

At the level of individual bacteria, we observed significantly increased abundances of *Haemophilus haemolyticus/influenzae, Corynebacterium macginleyi/accolens*, and *Fusobacterium* species in HCoV-positive cases compared to HCoV-positive controls. HCoV-positive cases also had amongst others significantly decreased abundances of *Moraxella* species, *Helcococcus* and *Corynebacterium propinquum/pseudodiphtheriticum* (Figure 1C). When comparing HCoV-negative cases to HCoV-negative controls, the differences in *Fusobacterium, C. macginleyi/accolens*, and *C. propinquum/pseudodiphtheriticum* abundance were not observed (Figure 1D). *H. influenzae/haemolyticus* was also significantly differentially abundant between HCoV-negative cases and HCoV-negative controls, though the effect size was smaller than between HCoV-positive cases and HCoV-positive controls. Importantly, we observed a trend toward increased *H. influenzae/haemolyticus* abundances in HCoV-positive cases (median abundance 32.7% [IQR 0.3-78.3]) when compared to HCoV-negative cases (median abundance 6.1% [IQR 0.1-49.4]; Wilcoxon test: p=0.080).

In summary, we have shown a high rate of 19.1% of asymptomatic HCoV carriage in healthy, community-dwelling children, which is in line with previous reports [7, 8]. HCoV carriage and disease seems, as most viral infections, highly seasonal in children. As aforementioned, mild and asymptomatic paediatric cases with high viral loads are also observed in the current COVID-19 pandemic [1–5]. The contribution to the spread of the disease by these children currently remains unknown, but is potentially substantial given significant viral shedding also occurs in asymptomatic children [6], and because they keep mixing with family members and other individuals in the community [12].

We have also shown that viral co-detection in HCoV-positive children differs between asymptomatic and symptomatic HCoV-positive children. Next to viral co-infection, increased abundance of several bacteria such as *H. influenzae/haemolyticus* and *Fusobacterium* is also associated with clinical symptoms of LRTI. By contrast, other bacteria like *Moraxella* as well as the presence of rhinovirus are associated with absence of symptoms. This implies that co-infection with specific viruses and bacteria may lead to HCoV-associated respiratory symptoms in young children. Regarding COVID-19, one study indeed found a viral co-detection rate of 40% for paediatric patients [13], while another study with predominantly adult patients found a much lower rate of 3.2% [14]. Whether microbial co-colonization may play an important role in developing (severe) symptomatology of COVID-19, in children and in adults, thus remains open to further investigation. This is however crucial to study to better understand disease pathogenesis, and thereby inform therapy and prevention of severe HCoV-associated infections.

## Data Availability

Sequence data that support the findings of this study have been deposited in the NCBI Sequence Read Archive (SRA) 189 database with BioProject ID PRJNA428382. Viral data are available from the corresponding author upon request.

https://www.ncbi.nlm.nih.gov/sra/?term=PRJNA428382

## Acknowledgements

We thank the participating children and their families. We also thank dr. Wing Ho Man (LUMC, Leiden, The Netherlands) and dr. Arine Vlieger (st. Antonius Hospital, Nieuwegein, The Netherlands) for their efforts in executing the study; the Bogaert lab for performing the laboratory work; and all the members of the research team at Spaarne Gasthuis Academy, the paediatricians at Spaarne Gasthuis, the Streeklaboratorium Haarlem, the GGD Kennemerland, and the JGZ Kennemerland for their support.

## References

1. Wei M, Yuan J, Liu Y, Fu T, Yu X, Zhang ZJ. Novel Coronavirus Infection in Hospitalized Infants under 1 Year of Age in China. JAMA 2020; in press.

2. Jiehao C, Jing X, Daojiong L, Zhi Y, Lei X, Zhenghai Q, Yuehua Z, Hua Z, Ran J, Pengcheng L, Xiangshi W, Yanling G, Aimei X, He T, Hailing C, Chuning W, Jingjing L, Jianshe W, Mei Z, Zeng M. A Case Series of children with 2019 novel coronavirus infection: clinical and epidemiological features. Clinical Infectious Diseases 2020; in press.

3. Liu W, Zhang Q, Chen J, Xiang R, Song H, Shu S, Chen L, Liang L, Zhou J, You L, Wu P, Zhang B, Lu Y, Xia L, Huang L, Yang Y, Liu F, Semple MG, Cowling BJ, Lan K, Sun Z, Yu H, Liu Y. Detection of Covid-19 in Children in Early January 2020 in Wuhan, China. New England Journal of Medicine 2020; in press.

4. Kam K, Fu Yung C, Cui L, Lin Tzer Pin R, Minn Mak T, Maiwald M, Li J, Yin Chong C, Nadua K, Woon Hui Tan N, Cheng Thoon K. A well infant with coronavirus disease 2019 (COVID-19) with high viral load. Clinical Infectious Diseases 2020; in press.

5. Tang A, Tong Z-D, Wang H-L, Dai Y-X, Li K-F, Liu J-N, Wu W-J, Yuan C, Yu M-L, Li P, Yan J-B. Detection of Novel Coronavirus by RT-PCR in Stool Specimen from Asymptomatic Child, China. Emerging infectious diseases 2020;26.

6. Xu Y, Li X, Zhu B, Liang H, Fang C, Gong Y, Guo Q, Sun X, Zhao D, Shen J, Zhang H, Liu H, Xia H, Tang J, Zhang K, Gong S. Characteristics of pediatric SARS-CoV-2 infection and potential evidence for persistent fecal viral shedding. Nature Medicine 2020; in press.

7. Heimdal I, Moe N, Krokstad S, Christensen A, Skanke LH, Nordbø SA, Døllner H. Human Coronavirus in Hospitalized Children With Respiratory Tract Infections: A 9Year Population-Based Study From Norway. The Journal of Infectious Diseases 2019; 219: 1198–1206.

8. Varghese L, Zachariah P, Vargas C, LaRussa P, Demmer RT, Furuya YE, Whittier S, Reed C, Stockwell MS, Saiman L. Epidemiology and Clinical Features of Human Coronaviruses in the Pediatric Population. Journal of the Pediatric Infectious Diseases Society 2018; 7: 151–158.

9. Man WH, van Houten MA, Mérelle ME, Vlieger AM, Chu MLJN, Jansen NJG, Sanders EAM, Bogaert D. Bacterial and viral respiratory tract microbiota and host characteristics in children with lower respiratory tract infections: a matched casecontrol study. The Lancet Respiratory Medicine 2019; 7: 417–426.

10. Pillet S, Lardeux M, Dina J, Grattard F, Verhoeven P, le Goff J, Vabret A, Pozzetto B. Comparative Evaluation of Six Commercialized Multiplex PCR Kits for the Diagnosis of Respiratory Infections. PLoS ONE 2013; 8: e72174.

11. Paulson JN, Stine OC, Bravo HC, Pop M. Differential abundance analysis for microbial marker-gene surveys. Nature Methods 2013; 10: 1200–1202.

12. Bi Q, Wu Y, Mei S, Ye C, Zou X, Zhang Z, Liu X, Wei L, Truelove SA, Zhang T, Gao W, Cheng C, Tang X, Wu X, Wu Y, Sun B, Huang S, Sun Y, Zhang J, Ma T, Lessler J, Feng T. Epidemiology and Transmission of COVID-19 in Shenzhen China: Analysis of 391 cases and 1,286 of their close contacts. medRxiv 2020; in press.

13. Xia W, Shao J, Guo Y, Peng X, Li Z, Hu D. Clinical and CT features in pediatric patients with COVID - 19 infection?: Different points from adults. Pediatric Pulmonology 2020; in press.

14. Lin D, Liu L, Zhang M, Hu Y, Yang Q, Guo J. Co-infections of SARS-CoV-2 with multiple common respiratory pathogens in infected patients. Science China Life Sciences 2020; 63.

